# Personalized Virus Load Curves of SARS-CoV-2 Infection

**DOI:** 10.1101/2021.01.21.21250268

**Authors:** Thomas Hillen, Carlos Contreras, Jay M. Newby

## Abstract

We introduce an explicit function that describes virus-load curves on a patient-specific level. This function is based on simple and intuitive model parameters. It allows virus load analysis without solving a full virus load dynamic model. We validate our model on data from influenza A as well as SARS-CoV-2 infection data for Macaque monkeys and humans. Further, we compare the virus load function to an established *target model* of virus dynamics, which shows an excellent fit. Our virus-load function offers a new way to analyse patient virus load data, and it can be used as input to higher level models for the physiological effects of a virus infection, for models of tissue damage, and to estimate patient risks.

---

Tracking the viral load in a patient’s body is critical in the treatment of viral infections. For many viruses, such as SARS-CoV-2, the progression of the viral load has a very typical time course, which can be classified into several temporal phases. Typically, an initial fast exponential increase leads to a virus load maximum, which is followed by a slow exponential decrease, followed by a fast exponential decrease leading to clearance (see Figure 1). Determining the duration and speed of these phases is important in the understanding of the disease progression in a given patient. Here we propose a simple model for the virus load that provides such biologically meaningful information. The model is based on simple, intuitive, model parameters, which can be directly related to clinical observations. The result is an explicit virus load function that can be used in higher level models which focus on the effect of a virus in the immune system, tissues and organs damage and person to person infectivity.

**Figure 1:**
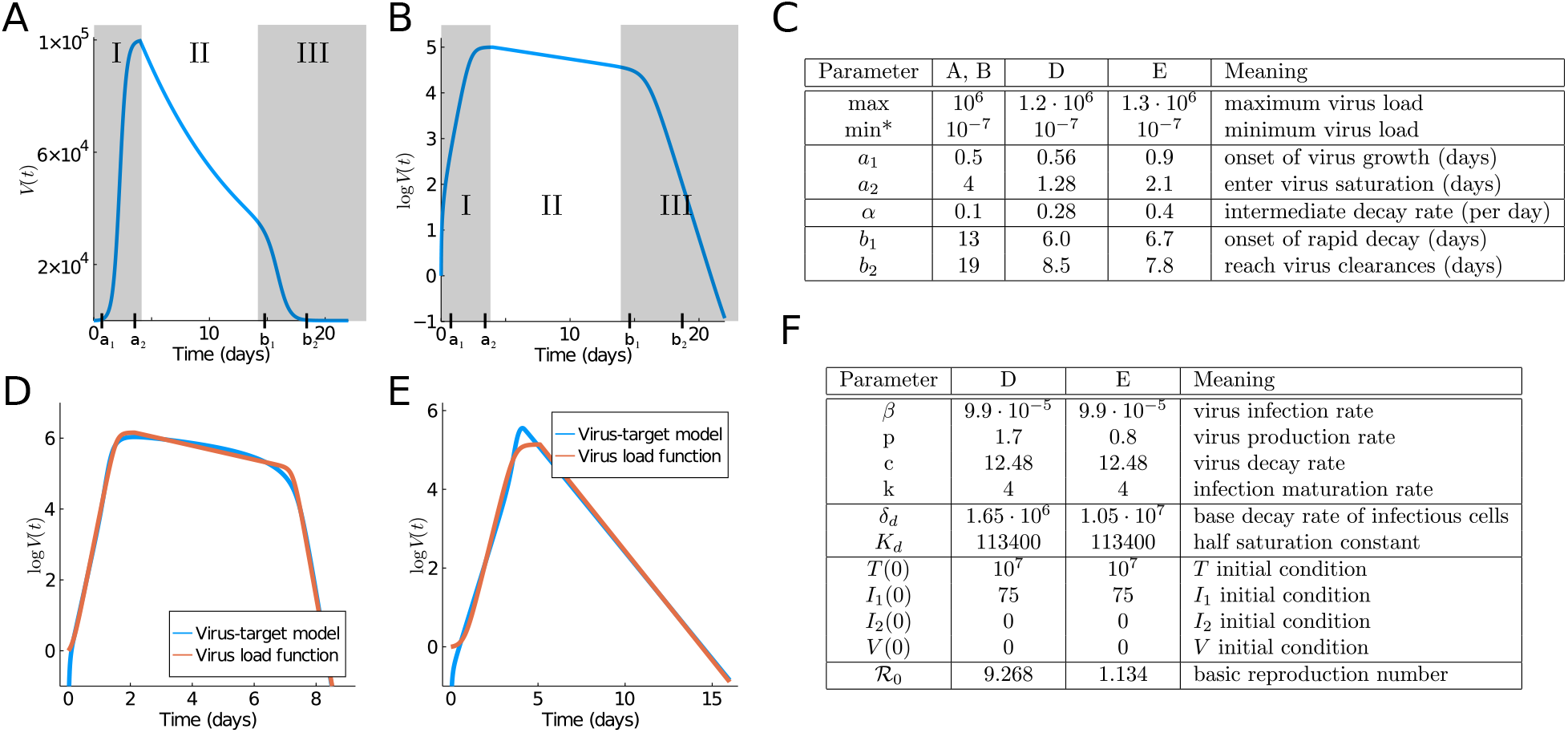
Typical virus load curves. The virus load (“titer”) is usually reported as a dilution value, TCID50, that is needed to infect 50% of a given cell culture in **(A)** absolute scale and **(B)** logarithmic scale. Shadow areas indicate the three phases in which we divide the virus load progression. **(C)** Parameters of the standard virus load function (1) corresponding to **A, B, D** and **E**. Virus load curve reported in [12] are used in **A** and **B. (D-E)** Comparison of the viral load curve from the viral target model (2) with the viral load function (1). Viral target curve showing triphasic and biphasic response are shown in **(D)** and **(E)**, respectively. Parameter values for the viral load function fitting to the viral target model are in **C. (F)** Parameters of the target model (2) corresponding to **D** and **E**.

Virus load curves, as reported in [12, 6, 5], have a very typical infection progression (see Figures 1**A** and **B**). In A. Smith [12] the virus infection has been classified into five phases, which we will combine into three phases for our purpose. In Smith’s classification, in the first phase (Phase Ia) the virus quickly infects cells without being detectable. This phase is followed by exponential growth (Phase Ib) until growth shows signs of saturation and a maximum is reached (Phase Ic). A period of slow exponential decline ensues, which we call Phase II. And finally, we often observe a fast decline that leads to virus clearance (Phase III). Depending on the virus and the response of the infected individual, these phases can be shorter or longer. The last Phase III is sometimes not seen in patient data, and the virus is cleared before the third phases starts. It is useful to distinguish between a tri-phasic behavior as in Figure 1**D** versus a bi-phasic behavior as in Figure 1**E**.

There is extensive use of ordinary differential equation models to describe virus-load curves [11, 2, 10]. For example Baccam *et al*. [1] developed a four-compartment virus-target model to describe the virus load in a given person. A. Smith [12] extended this model to include a viral clearance saturation term, which allowed her to fit mouse viral infection data. We will discuss this target model later in the Results section when we compare it to our virus load function.

To develop our virus load function below (see (1)) we consider the Phase I of sigmoid increase between time points *a*_1_ and *a*_2_ (see Figures 1**A** and **B**). The Phase I includes the three initial phases (Phases Ia, Ib, Ic) of Smith [12] mentioned above. At time *a*_2_ a slow decline of the virus is observed as the immune response kicks in (Phase II between *a*_2_ and *b*_1_ in Figure 1) and finally (Phase III) shows a rather sharp decline once the virus is controlled (between *b*_1_ and *b*_2_). We write the virus load curve as a product of three functions, representing the three main phases:

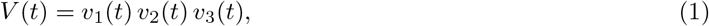

where *v*_1_ describes the initial growth phase between *a*_1_ and *a*_2_, *v*_2_ the intermediate slow decay phase between *a*_2_ and *b*_1_, and *v*_3_ the final decay phase between *b*_1_ and *b*_2_. These are given as sigmoid and exponential functions, respectively

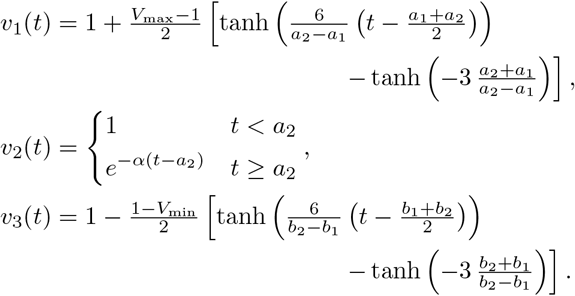

The specific form of sigmoid curves for *v*_1_ and *v*_3_ was developed previously by Olobatuyi in [7] in a cancer model and more details are given in *Materials and Methods*. It allows us to define these functions based on intuitive transition threshold values. The value *a*_1_ describes the onset of growth and *a*_2_ a value when saturation is reached, similarly, *b*_1_ denotes the time where decay switches from slow to fast and *b*_2_ is the time when the virus is effectively eliminated. The parameter *α* describes the intermediate exponential decay rate. In Figure 1**C** we list the values used in Figures 1**A** and **B** and their meaning. This virus load function can describe the tri-phasic or bi-phasic response, as shown in Figure 1**D** and **E**.

Virus load functions are in high demand in the virus modelling community. For example in [4], a large community of researchers develops an individulal based SARS-CoV-2 physiological model that includes virus infection, virus transmission, immune response, and potential damage to the tissue. The immune response and the tissue complications are directly related to the virus load of the tissue. Our standard virus load function will be a welcome modelling tool to describe tissue damage and assess complication risks. Another example of detailed virus modelling is a recent preprint on the impact of SARS-CoV-2 on the renin-angiotensin-system by Pucci et al. [8]. A realistic virus load function is needed as model input in their model. The pharmacological company Pfizer made it their emphasis to develop new treatment strategies and new estimates of side effects, once a Covid-19 treatment becomes available. A virus load function, as presented here, will be a welcome tool to test their ideas.^1^

## Results

We fit (1) to virus load data of infection of influenza A of mice, and to SARS-CoV-2 data for humans and Macaque monkeys. Details on these data sets and our data fitting procedure it explained in *Materials and Methods*. Furthermore, we compare this new virus load function (1) to the standard target model for viral kinetics [12].

### Influenza virus load data

We begin with Influenza A virus load data from [12] as these are the best experimental data available, based on tightly controlled murine experiments. In [12] ten mice were infected with influenza A virus and time series of virus load titers on the log-scale were measured. Initial conditions for the virus-target model are also available in that reference. In Figure 2, we show those data plus the data fitting results of the viral load function (1) and the viral target model 2, which we discuss in detail later. The fit of the target model to these data was originally performed by Smith *et al*. in [12]. Of particular importance is the virus-load decay rate in Phase II. Our virus load function estimates the negative slope as *α* =− 0.26 d^*−*1^, quite similar to the estimate in [12] using linear regression for the middle portion (−0.2 d^*−*1^). The virus load function estimates the duration of Phase I at approximately 2.41 days, Phase II at 3.22, days and Phase III at 1.35 days.

**Figure 2:**
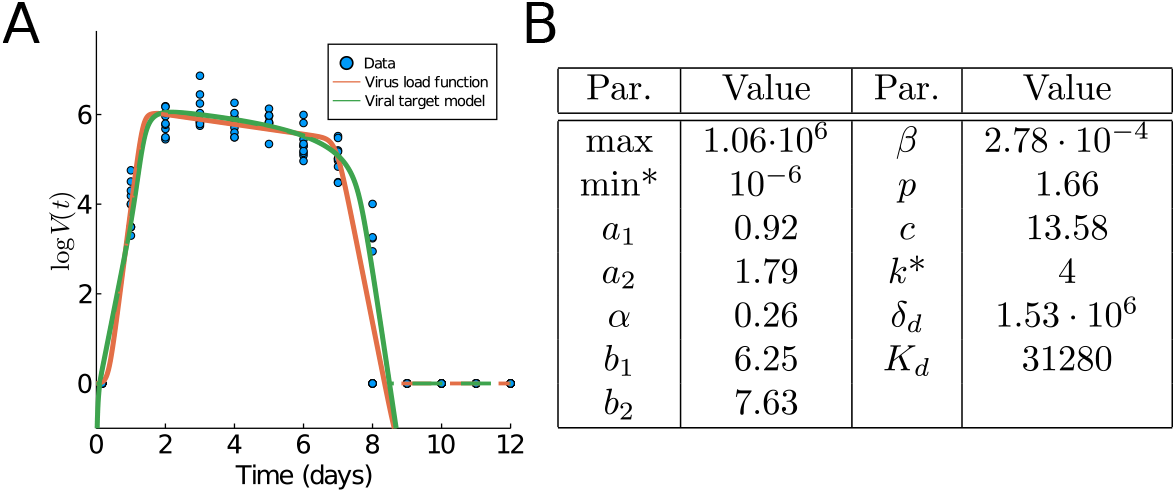
Fitting results of the virus load function (1) and (2) to influenza A data [12]. **(A)** Graph with data and best fit for both models. The empirical virus load curves are indicated with dashed lines. **(B)** Best estimate for the parameters. The best estimate for the viral target model were directly communicated by one of the authors of [12]. The initial conditions are the same as in Figure 1**F**. Fixed parameters are denoted with ^*∗*^.

### Human SARS-CoV-2

In [13] a cohort of COVID-19 patients from two Hong Kong hospitals was evaluated. 30 patients were screened between Jan. 22, 2020 and Feb 12, 2020, and 23 patients were included in the study (see Figure 5).

For each patient on a daily basis a multitude of clinical measurements were recorded, including a virus-load measurement. For patients who were not intubated an oropharynx saliva sample was collected. In the early mornings patients were asked to cough up to clear the throat and the virus load in the saliva was measured. From patients who were intubated a endotrachial aspirate sample was taken. As the ciliary activity of the lung epithelium transports mucus to the posterior oropharyngeal area, these samples give a good indication of the viral activity in the lungs. The data were collected on a daily basis and recorded as mean values and standard deviations. We fit our virus load function (1) to eight (8) patients in this data set as shown in Figures 3**A**–**H**, and the corresponding model parameters are listed in Figure 3**I**.

**Figure 3:**
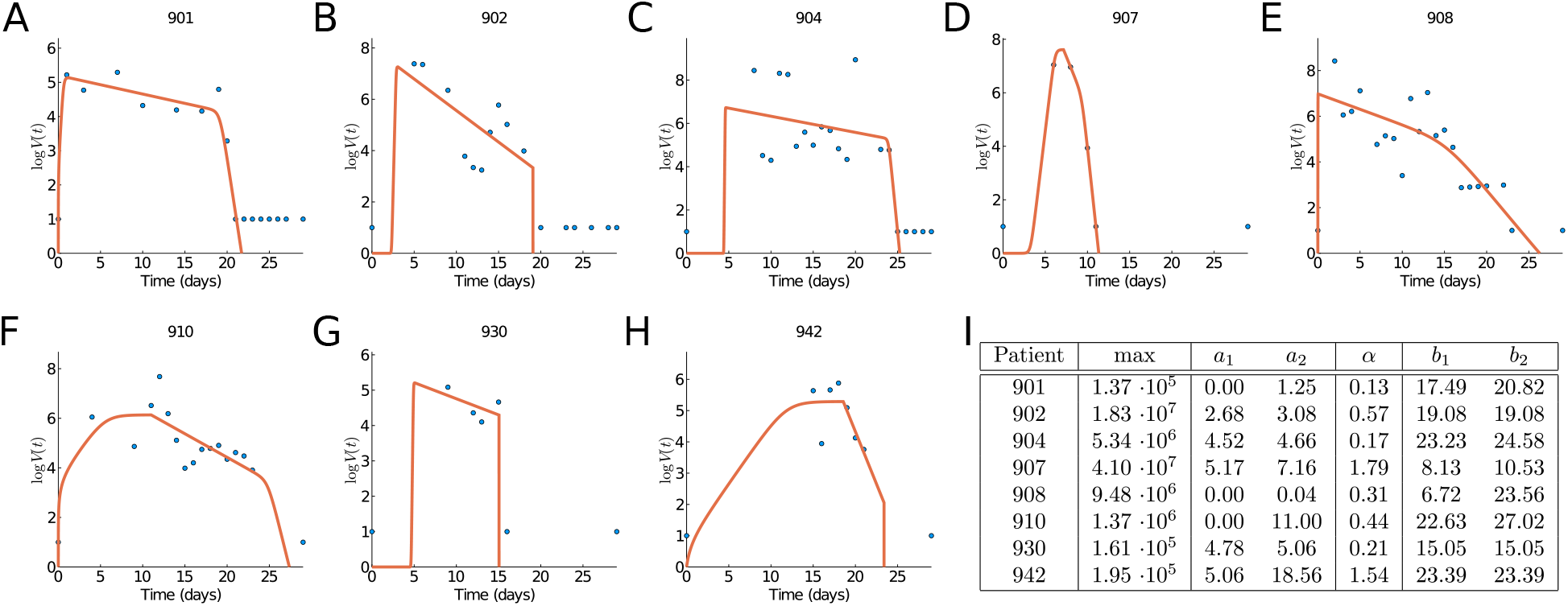
Fitting results of the virus load function (1) to data. **(A-H)** Fitting to patients 901, 902, 904, 907, 908, 910, 930, and 940 from [13]. **(I)** Estimated parameter values.

We observe that the virus load over time varies greatly across patients, some show long infection periods (20-25 days), others are short (∼ 10 days). The virus load function is able to describe the three phases of the virus for most of the patients (901, 902, 904, 908, and 930). In those patients, we observe the initial virus growth phase is rather quick and the virus reaches its carrying capacity within a day (*a*_2_−*a*_1_ *<* 1.08). The slope during the second phase varies from -0.13 to -0.57. The virus load reaches a saturation level, which is likely to be related to the innate immune response and it starts a phase of slow decay with a half-life time between 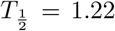 and 5.33 days. After 15 days the virus load drops more quickly, possibly due to the adaptive immune response and at day 25 the virus is cleared. Note that this is sensitive to missing data and drastic changes during both phases. For instance, in patients 902, 904, 907, 930, and 942 there is no data at the beginning for several days. Note also that, for example, for patient 930 there is an sudden full clearance of the virus at day 15. As consequence, *b*_1_ and *b*_2_ are estimated as equal. For patients 907 the fit is exact, but it should be noted that in this case we have more parameters than data points, and a good fit is not very meaningful. For patient 930 we also have a very few data points, but they seem to be more scattered to obtain an exact fit. Patients 930 and 942 show an extended initial phase I which spans over 11 days patient 910 and 13 days for patient 942. This is, of course, related to the missing initial information about the early infection times.

We see that the virus load function (1) describes all virus load curves very well. It is easily adapted for long and short virus infection periods and it is fully flexible in detecting all three phases of the viral progression. We will see, the dynamics for macaque monkeys is rather similar.

### Macaque Monkey Data

In [3] nine rhesus macaques monkeys were infected with SARS-CoV-2. Three monkeys (group 1) obtained a high initial virus dose, three monkeys (group 2) a medium initial dose, and three (group 3) a small initial dose. The virus load was measured daily or every other day through a bronchoalveolar lavage probe. The recorded data for each individual group and for all three groups together are shown as dots in Figure 5. All nine monkeys showed only mild disease symptoms and they all fully recovered. Hence the infection cycle here is more indicative of a mild infection, in contrast to the human data considered above.

In Figures 4**A**–**I** we fit our virus load function (1) to the rhesus monkey data, and parameter values are in Figure 4**J**. The characteristic values for the initial virus growth *a*_2_ *<* 2.3 is common between all monkeys, indicating that the amount of initial viral dose is not so important. The virus load is biphasic in most monkeys (1-1, 1-2, 1-3, 2-2, 2-3, and 3-2) in which the rate of decay is larger (*>* 0.6 days^*−*1^, and half-life time *<* 1.2 days). For the remaining monkeys (2-1, 3-1, and 3-3) a fast decay phase is observed, following a slow decay with smaller decay rate. This distinction between biphasic and triphasic viral load suggests that in some monkeys the action of the immune system is rather efficient.

**Figure 4:**
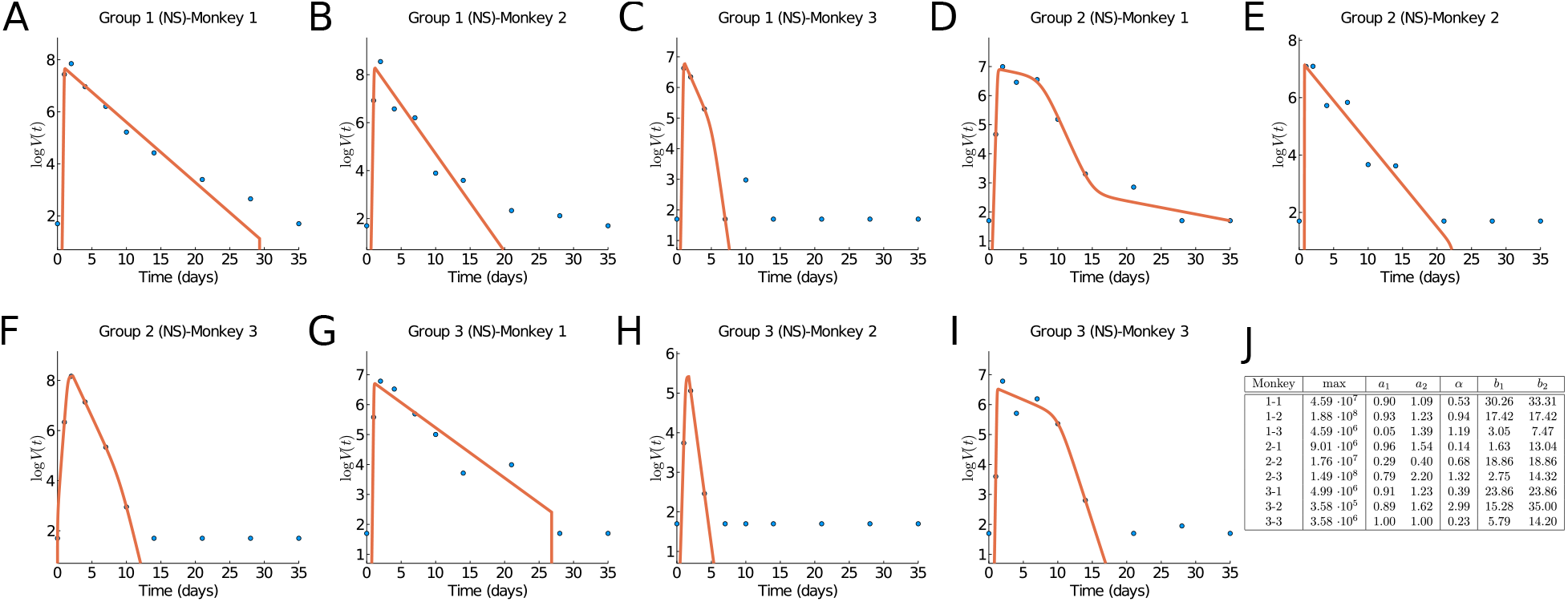
Results of fitting the virus load function (1). **(A-I)** Groups 1, 2, and 3, monkeys 1, 2, and 3. Data from [3]. **(J)** Estimate parameter values.

**Figure 5:**
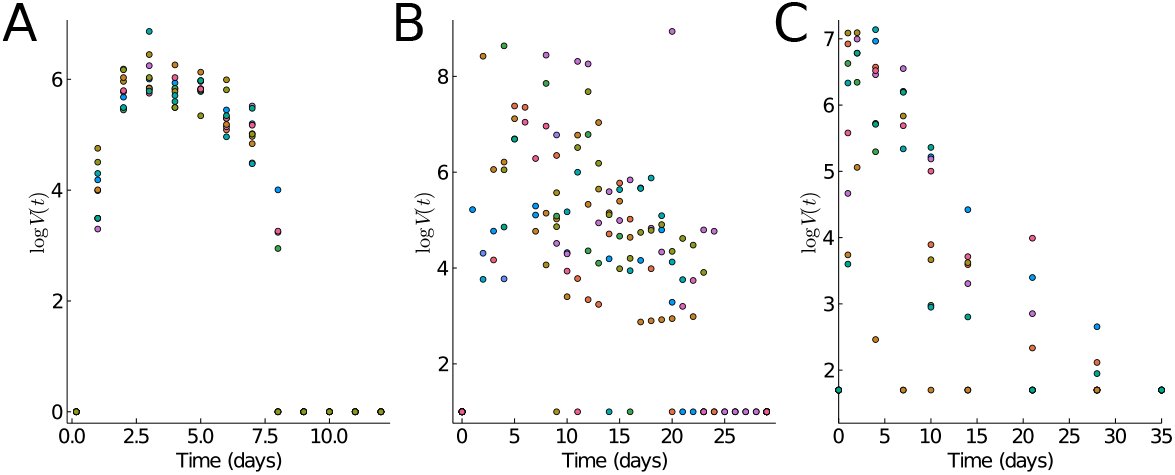
**(A)** Mice Influenza A data [12]. **(B)** Human SARS-Cov2 data from oropharynx saliva samples [13]. **(C)** Macaque monkey SARS-Cov2 [3].

Compared to the human data we notice that the virus half-life times in Phase II is 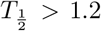 days in both human and monkeys experiencing all three phases, and 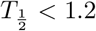 days in those which show a biphasic behavior. Hence a biphasic behavior is indicative of faster virus clearance in Phase II. The length of the infection is estimated as about 27.5 days and for humans and 34 days for the monkeys, where the final decay phase starts significantly earlier in humans *b*_1_ = 15 than in monkeys *b*_1_ = 25−30. This could be an indication of a more efficient adaptive immune response in humans as compared to monkeys.

### Comparison to the Virus-target model of Smith et al

As mentioned in the *Introduction*, the virus load modelling with ordinary differential equations is a well developed field [11, 2, 10]. Here we like to compare our virus load function (1) with one of the most current target models, the model of A. Smith *et al*. [12]. The model of A. Smith [12] is based on an earlier model of Baccam [1] and it is modified so it can explain all three Phases as well. This virus load model has been fit in [12] to the influenza data mentioned above and we show this fit as a green line in Figure 2 **A**. The so called target model comprises of four ordinary differential equations (ODEs) for the target cells *T* (*t*), the infected cells, also called the eclipse phase, *I*_1_ (*t*), the infectious cells *I*_2_ (*t*) and the virus load *V* (*t*),

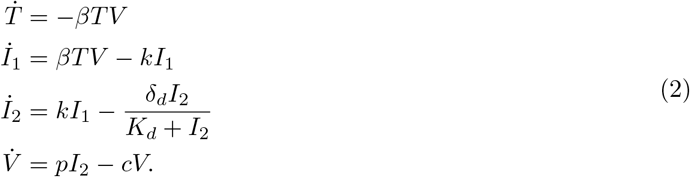

Here *β* is the virus infection rate, *k* the infection maturation rate, *p* the virus production rate, *c* the virus decay rate, *δ*_*d*_ the base decay rate of infectious cells, and *K*_*d*_ the half saturation constant for the decay term of the infectious cells. This model is an improvement of the standard viral kinetic model [1] in which the authors introduce a saturation term for the infected cell clearance 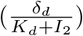 in the equation for *I*_2_ to describe a triphasic virus growth and decay [12]. The basic reproduction number *R*_0_ for this model is given as (see [12])

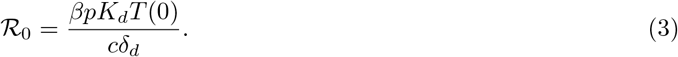

In Figure 1**D** and **E** we show the virus load curve from the target model as blue line showing a triphasic and biphasic type, respectively, in virus growth and decay using parameters values listed in **F**. Note that the corresponding *ℛ*_0_ values (3) are significantly different and higher for the triphasic case. To these curves, we fit the virus load function (1) as red curves (see *Materials and Methods* for details). We see that the virus load function (1) can reproduce both biphasic and triphasic curves with a high level of accuracy. The best estimate for the parameter values are listed in Figure 1**C**.

Fitting the virus-target model (2) to data constitutes statistical and numerical challenges. For instance, initial conditions are unknown in must cases, and estimating these values has shown to be impractical [12]. However, as shown below, it is easier to fit the virus load function (1) to virus load data due to the empirical nature of the parameter in the function.

## Conclusion

The explicit form of this new standard virus load function (1) is simple and convenient. The corresponding parameter values all have easily understandable biological meaning, allowing this model to be used quickly and efficiently.

We have shown that this virus load function can replicate observed virus load titers from Influenza A in mice and from SARS-CoV-2 in humans and in monkeys. A quick analysis shows already that macaque is a good model system for human infection. The early and intermediate infection phases are very similar, and the final elimination period is quicker in humans.

It should be noted that estimating *a*_1_, *a*_2_, *b*_1_, and *b*_2_ require further consideration. For instance, for patient 930 Figure 3**G**, it is easy to see from the graph that any values 0 *< a*_1_ *< a*_2_ *<* 9 are good estimations. This is because of missing data in the first phase leading to a large uncertainty about the start and the duration of the outbreak of the virus in that particular patient.

There are numerous potential uses of the this virus load function (1). It can be used as input into further models for tissue damage through a virus infection such as those presented in [4] on lung tissue modelling or on the angiostatin pathway. Once more human data are available it can be used to perform population statistics on infection period length and virus decay rates, and it will enable us to estimate patient risk. Based on the parameters, it might be possible to classify individuals as fast versus slow responders or low versus high risk patients. All this work is ongoing, and having a simple tool as presented here will be invaluable.

## Materials and Methods

### Data fitting procedure

We fit the viral load function (1) to three sets of data shown in Figure 5: mice influenza A from [12], human SARS-CoV-2 data from [13], and Macaque monkeys SARS-Cov-2 from [3]. Unfortunately, we do not have sex or gender information for any of these data. The virus load titer is measured as a relative RNA expression as compared to a reference gene. Hence the measurements have a significant measurement threshold *θ* and virus loads below this threshold cannot be seen. This threshold is 0 (on a logarithmic scale) for the Influenza A data above, 1 for the human data above, and 1.7 for the Macaque monkey data. Hence measurement values at the threshold cannot be used for the fitting of the curves, since the virus load might be lower than recorded. To account for this, we fit the data to the *effective* viral load function

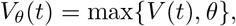

where *θ* is the detection threshold given by the data. This explains why the fitted curves in Figures 3 and 4 seem to “ignore” the non-detection values that are shown for large times.

For the SARS-Cov-2 data, we fit the viral load function to each subject separately since each subject seems to experience a difference response for these two data sets. We also assume that each subject starts and ends with viral load equal to the threshold. Hence we force those values if necessary. We also ignore subjects with less than 5 observations.

To fit the data we use the Levenberg-Marquard algorithm available in the *LsqFit*.*jl* package for Julia. The initial guess for *a*_1_, *a*_2_, *b*_1_, and *b*_2_ is taken from an ordered sample of four times uniformly distributed between the 0 and the maximum time. The lower and upper bounds are set to be evenly spaced around those initial guess. The initial guess for max is taken as the maximum data ±3 to define the bounds. The value of min = −6 is fixed. Finally, the initial guess for *α* is 0.8 bounded between [10^*−*8^, 10^4^]. We found the best fit by starting from ten thousand random initial guesses and choosing the lowest residual sum of squares.

### Hyperbolic Tangent

The hyperbolic tangent function,

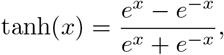

is a sigmoid step function that smoothly transitions from −1 to 1. In (1) it has been shifted and scaled such that transitions occur between *a*_1_ and *a*_2_ upwards and between *b*_1_ and *b*_2_ downwards, where the maximum is *max* and the minimum *min*. The value “6” in the argument of the hyperbolic tangent can easily be explained by looking at *a*_1_ = −1 and *a*_2_ = 1. Then tanh 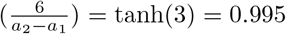. That means that at *a*_2_ a 99.5% saturation level is reached. Similar, at *a*_1_ the function is 0.5% above its minimum.

## Data Availability

data is available by request to the authors of references 1, 11, and 12

## Footnotes

1 Pfizer has, due to proprietary reasons, not yet published on Covid-19 modelling. My statements above are based on personal communications.

